# Vitamin D deficiency as risk factor for severe COVID-19: a convergence of two pandemics

**DOI:** 10.1101/2020.05.01.20079376

**Authors:** D. De Smet, K. De Smet, P. Herroelen, S. Gryspeerdt, G.A. Martens

## Abstract

**Context:** Through its immunological functions, vitamin D attenuates inflammatory responses to respiratory viruses. Vitamin D deficiency might be a highly prevalent risk factor for severe SARS-CoV-2 infections.

**Objective:** To investigate the level of vitamin D deficiency in West Flanders, Belgium and its correlation to severity of COVID-19 as staged by CT

**Design:** Retrospective observational study

Setting. Central network hospital

**Participants:** 186 SARS-CoV-2-infected patients hospitalized from March 1, 2020 to April 7, 2020

**Main outcome measure:** Analysis of 25(OH)D in COVID-19 patients versus season/age/sex-matched diseased controls

**Results:** The rate of vitamin D deficiency (25(OH)D<20 ng/mL) in West Flanders varies with age, sex and season but is overall very high (39.9%) based on analysis of 16274 control samples. We measured 25(OH)D levels in 186 COVID-19 patients (109 males (median age 68 years, IQR 53-79) and 77 females (median age 71 years, IQR 65-74)) and 2717 age/season-matched controls (999 males (median age 69 years, IQR 53-81) and 1718 females (median age 68 years, IQR 43-83)). COVID-19 patients showed lower median 25(OH)D (18.6 ng/mL, IQR 12.6-25.3, versus 21.5 ng/mL, IQR 13.9-30.8; P=0.0016) and higher vitamin D deficiency rates (58.6% versus 45.2%, P=0.0005). Surprisingly, this difference was restricted to male COVID-19 patients who had markedly higher deficiency rates than male controls (67.0% versus 49.2%, P=0.0006) that increased with advancing radiological stage and were not confounded vitamin D-impacted comorbidities.

**Conclusions:** vitamin D deficiency is a prevalent risk factor for severe COVID-19. Vitamin D supplementation might be an inexpensive and safe mitigation for the SARS-CoV-2 pandemic.

SUMMARY BOX
What is already known on this topic
Vitamin D deficiency increases the incidence and severity of respiratory viral infections by exacerbation of pro-inflammatory immune responses. Lung damage in COVID-19 involves excessive inflammation and cytokine release. With more than 1 billion people worldwide affected by vitamin D deficiency the world might now face a convergence of two pandemics. Data are emerging that variations in vitamin D deficiency rates across ethnic and demographic subgroups are correlated to severity of SARS-CoV-2 infections.
What this study adds
Our data reveal that more than 40% of the adult population in a wealthy European country is vitamin D deficient. We show that vitamin D deficiency is correlated with the risk for hospitalization for COVID-19 pneumonia and predisposes to more advanced radiological disease stages. Specifically, men were at risk. This correlation was not confounded by vitamin D-impacted morbidities such as coronary artery disease, diabetes and chronic lung disease. Our findings support a causal relation between vitamin D deficiency and severe COVID-19 and call for vitamin D supplementation as safe, widely available and inexpensive mitigation strategy.

## Introduction

In severe SARS-CoV-2 infections excessive activity of pro-inflammatory immune cells contributes to alveolar and endothelial damage triggering a vicious cycle that evolves towards severe COVID-19 ^1,2^. Beside its role in calcium metabolism, 1,25-dihydroxyvitamin D (25(OH)D) is a pleiotropic regulator of the immune system ^3,4^. It stimulates the expression of cathelicidins and beta-defensin in respiratory epithelia as barrier to pathogen invasion ^5,6^. It acts as a pro-tolerogenic cytokine dampening excessive inflammation by inhibiting neutrophils and switching Th1 CD4 T cells and M1-polarized macrophages towards a type II immunity. Vitamin D deficiency increases the severity of respiratory virus infections ^7,8^ and contributes to variations in their incidence across seasons, age groups, socioeconomic status and geographies. Data are emerging that vitamin D deficiency is more prevalent in patients with severe COVID-19 pneumonia requiring intensive care ^9^. More than a billion people worldwide are vitamin D deficient^10^ with variations between sexes, ethnicities, social groups and geographies that appear to correlate with differences in incidence and outcome of COVID-19 lung disease based on modeling of large data sets ^11-13^. Here we provide a first field validation of these models. We provide a detailed view on the surprisingly high prevalence of vitamin D deficiency in a wealthy European region, the province of West Flanders, Belgium. We investigated the correlation of serum 25-hydroxyvitamin D (25(OH)D) level to the risk to be hospitalized for severe COVID-19 pneumonia, the radiological disease stage as proxy for immunological stage and its possible confounding by vitamin D-impacted comorbidities, in cohort of 186 consecutive patients hospitalized for severe SARS-CoV-2 infection in a large Belgian network hospital.

## Methods

### Patients

This is a retrospective observational study on 186 consecutive patients hospitalized from March 1, 2020 to April 7,2020 for COVID-19 pneumonia at AZ Delta General Hospital in Roeselare, Belgium (demographics in Table 2). 25(OH)D levels in COVID-19 patients were compared to an age- and season-matched diseased control population, consisting of 2717 consecutive unselected patients sampled from March 1, 2019 to April 30, 2019. The prevalence and age/sex/seasonal-distribution of vitamin D status in the general population was derived from all 25(OH)D measurements from January 1, 2019 to December 31, 2019 on 16274 consecutive, unselected and unique patient samples (Table 1). This study was approved by the AZ Delta ethical committee (Clinical Trial Number IRB B1172020000009) with a waiver of informed consent from study participants considering the study is based on secondary analysis of existing data.

**Table 2:**
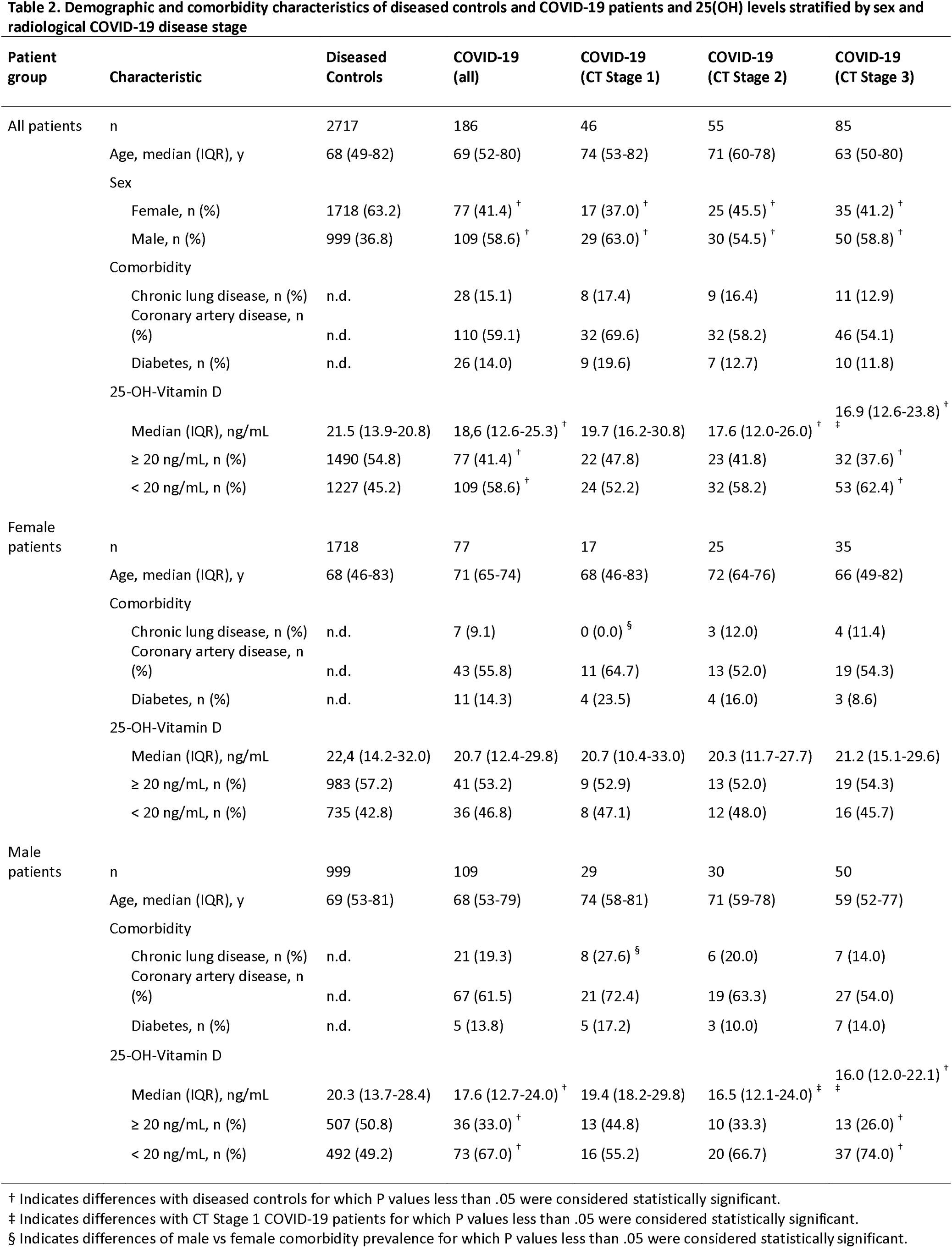
Demographic and comorbidity characteristics of diseased controls and COVID-19 patients and 25(OH) levels stratified by sex and radiological COVID-19 disease stage. † Indicates differences with diseased controls for which P values less than .05 were considered statistically significant. % Indicates differences with CT Stage 1 COVID-19 patients for which P values less than .05 were considered statistically significant. § Indicates differences of male versus female comorbidity prevalence for which P values less than .05 were considered statistically significant. Data (not normally distributed) are expressed as medians (25th–75th percentiles), and the Mann-Whitney test was used to test statistical difference between groups. Proportions for categorical variables were compared using chi-squared test. Exact P values listed in Supplementary information.

**Table 1:** Sex, age and seasonal variations in 25(OH) levels in the general population as derived from 25(OH)D measurements in 16274 unselected, consecutive and unique diseased control samples. The table lists for the number (percentage) of patients for the indicated groups, median (IQR) age (years), median (IQR) 25(OH)D level (ng/mL) and percentage vitamin D deficiency. Data (not normally distributed) are expressed as medians (25th–75th percentiles), and the Mann-Whitney test was used to test statistical difference between groups. Proportions for categorical variables were compared using chi-squared test. Upper case letters indicate relevant statistical differences detailed in the footnote.

| Patient group | n (%) | Age, Median (IQR), y | 25-OH-Vit D, Median (IQR), ng/mL | 25-OH-Vit D < 20 ng/mL, n (%) |
| --- | --- | --- | --- | --- |
| **Gender distribution** |  |  |  |  |
| All | 16274 (100) | 64.2 (39.8-81.3) | 23.3 (15.3-32.4) | 6491 (39.9) |
| Female | 10045 (61.7) | 63.3 (37.4-82.0) | 23.7 (15.6-33.1) | 3875 (38.6) |
| Male | 6229 (38.3) | 65.2 (44.5-80.3) | 22.3 (14.9-31.3)<sup>a</sup> | 2616 (42.0)<sup>b</sup> |
| **Age (year) distribution** |  |  |  |  |
| **All** |  |  |  |  |
| ≤ 1 | 113 (0.7) | 0.7 (0.3-0.8) | 44.4 (37.3-55.1) | 6 (5.3) |
| 1-10 | 761 (4.7) | 5.0 (2.9-7.3) | 32.7 (26.0-42.0) | 83 (10.9) |
| 10-18 | 399 (2.5) | 13.1 (11.5-15.3) | 23.9 (17.4-30.0) | 130 (32.6) |
| 18-30 | 1410 (8.7) | 25.8 (22.6-27.9) | 22.2 (15.5-30.0)<sup>c, d</sup> | 600 (42.6)<sup>e, f</sup> |
| 30-50 | 2842 (17.5) | 40.1 (34.9-45.5) | 22.5 (15.7-30.6)<sup>c, d</sup> | 1171 (41.2)<sup>e</sup> |
| 50-70 | 3727 (22.9) | 60.1 (55.4-65.1) | 23.4 (15.6-32.6)<sup>c</sup> | 1467 (39.4)<sup>e</sup> |
| >70 | 7022 (43.1) | 82.9 (77.5-87.7) | 22.3 (13.9-31.9)<sup>c, d</sup> | 3034 (43.2)<sup>e, f</sup> |
| **Female** |  |  |  |  |
| ≤ 1 | 49 (0.5) | 0.7 (0.4-0.8) | 44.8 (38.8-54.6) | 4 (8.2) |
| 1-10 | 360 (3.6) | 5.5 (3.2-7.3) | 32.9 (25.5-41.7) | 33 (9.2) |
| 10-18 | 240 (2.4) | 13.6 (11.5-15.5) | 23.3 (17.0-29.9)<sup>g</sup> | 83 (34.6)<sup>i</sup> |
| 18-30 | 1088 (10.8) | 26.2 (23.1-28.0) | 22.4 (15.6-30.1)<sup>g, h</sup> | 455 (41.8)<sup>i, j</sup> |
| 30-50 | 1862 (18.5) | 39.1 (34.3-44.6) | 22.9 (15.9-31.6)<sup>g, h</sup> | 737 (39.6)<sup>i</sup> |
| 50-70 | 2104 (20.9) | 59.7 (55.2-64.8) | 24.7 (16.5-34.2)<sup>g</sup> | 766 (36.4)<sup>i, j</sup> |
| >70 | 4342 (43.2) | 83.6 (78.2-88.1) | 23.1 (14.0-32.9)<sup>g, h</sup> | 1797 (41.4)<sup>i</sup> |
| **Male** |  |  |  |  |
| ≤ 1 | 64 (1.0) | 0.6 (0.3-0.8) | 44.3 (37.1-55.7) | 2 (3.1) |
| 1-10 | 401 (6.4) | 4.7 (2.6-7.3) | 32.1 (26.3-42.0) | 50 (12.5) |
| 10-18 | 159 (2.6) | 12.8 (11.4-14.9) | 24.5 (18.7-31.1) | 47 (29.6) |
| 18-30 | 322 (5.2) | 24.4 (21.2-27.4) | 21.6 (14.9-29.1)<sup>k</sup> | 145 (45.0)<sup>m</sup> |
| 30-50 | 980 (15.7) | 41.7 (36.4-46.2) | 21.8 (15.5-29.3)<sup>k, l</sup> | 434 (44.3)<sup>m, n</sup> |
| 50-70 | 1623 (26.1) | 60.7 (55.6-65.5) | 21.9 (14.4-30.7)<sup>k, l</sup> | 701 (43.2)<sup>m, n</sup> |
| >70 | 2680 (43.0) | 81.9 (76.5-86.7) | 21.2 (13.8-30.1)<sup>k, l</sup> | 1237 (46.2)<sup>m, n</sup> |
| **Seasonal distribution** |  |  |  |  |
| **Winter** |  |  |  |  |
| All | 3889 (100) | 62.5 (37.7-81.4) | 21.8 (14.3-31.4)<sup>o</sup> | 1740 (44.7)<sup>q</sup> |
| Female | 2448 (62.9) | 62.5 (36.1-81.3) | 22.6 (14.5-32.4)<sup>o</sup> | 1040 (42.5)<sup>q</sup> |
| Male | 1441 (37.1) | 62.8 (40.9-79.5) | 20.4 (14.1-29.8)<sup>o, p</sup> | 700 (48.6)<sup>q, r</sup> |
| **Spring** |  |  |  |  |
| All | 4277 (100) | 65.8 (40.5-81.6) | 22.2 (14.4-31.0)<sup>o</sup> | 1832 (42.8)<sup>q</sup> |
| Female | 2563 (59.9) | 64.2 (37.3-82.0) | 22.4 (14.8-31.6)<sup>o</sup> | 1075 (41.9)<sup>q</sup> |
| Male | 1714 (40.1) | 67.5 (46.3-81.0) | 21.8 (14.0-30.3)<sup>o, p</sup> | 757 (44.2)<sup>q</sup> |
| **Summer** |  |  |  |  |
| All | 3619 (100) | 63.3 (40.6-81.6) | 25.7 (17.0-35.0) | 1202 (33.2) |
| Female | 2296 (63.4) | 63.6 (38.8-82.5) | 25.8 (17.2-35.4) | 748 (32.6) |
| Male | 1323 (36.6) | 62.9 (43.3-80.1) | 25.6 (16.7-34.2) | 454 (34.3) |
| **Fall** |  |  |  |  |
| All | 4489 (100) | 64.7 (40.1-80.9) | 23.6 (15.9-32.6)<sup>o</sup> | 1717 (38.3)<sup>q</sup> |
| Female | 2738 (61.0) | 63.1 (37.4-81.3) | 24.2 (16.1-33.3)<sup>o</sup> | 1012 (37.0)<sup>q</sup> |
| Male | 1751 (39.0) | 66.3 (45.7-80.3) | 23.0 (15.6-31.3)<sup>o,p</sup> | 705 (40.3)<sup>q,r</sup> |

### Procedures

*Chest CT* On admission all COVID-19 patients received a chest CT (detailed CT scanning protocol in Supplementary Information) to determine the disease stage by consensus evaluation of the predominant radiological presentation: ground-glass opacities (early stage, 0-4 days, “stage 1”), crazy paving pattern (progressive stage, 5-8 days, “stage 2”), (3) consolidation (peak stage, 10-13 days, “stage 3”)^14^. *Analysis of comorbidities:* prevalence of diabetes was registered by anamnesis and review of electronic patient records. Chronic lung disease (emphysema, fibrosis, bronchiectasis) and coronary artery disease (coronary artery calcification scoring) were objectified by chest CT. *Laboratory analyses:* all serum 25(OH)D measurements in this study were done in a central lab by the exact same method using Elecsys® vitamin D total II (Roche, Switzerland) traced to the official reference ID-LC-MS/MS (Ghent University). SARS-CoV-2 infection was confirmed in all COVID-19 patients by PCR for E/N/RdRP genes (Allplex™ 2019-nCoV assay, Seegene, Seoul, Korea) on nasopharyngeal swabs.

### Statistical analysis

Data (not normally distributed) are expressed as medians (IQR) and Mann-Whitney test was used to test statistical differences between groups. Proportions for categorical variables were compared using chi-squared test. Spearman’s rank correlation was used to identify correlations between 25(OH)D and comorbidities as confounding factors (r_s_, 95% CI). Statistical analyses were performed using MedCalc (version 12.2.1, Mariakerke, Belgium) and considered significant if P value was less than .05.

## Results

### Epidemiology and distribution of vitamin D deficiency in the general population

The prevalence and distribution of vitamin D deficiency the general population in the province of West Flanders, Belgium, was derived from 25(OH)D measurements in 16274 consecutive, unselected and unique patient sampled from January 1, 2019 to December 31, 2019. Table 1 summarizes the demographics and distributions according to sex, age and season and lists relevant statistical differences. Males had lower median 25(OH)D than females (22.3 ng/mL, IQR 14.9-31.3 versus 23.7 ng/mL, IQR 15.6-33.1, P<0.0001) and higher rates of vitamin D deficiency (42.0% versus 38.6%, P<0.0001) (Fig. 1A). With the exception of the very young (<10 years), vitamin D deficiency was endemic in all age groups (Fig. 1B, Table 1). Men and women above 18 years showed lower median 25(OH)D levels and higher vitamin D deficiency rates than individuals of 18 years or younger (P<0.05). As expected, 25(OH)D levels were lower in winter and spring than in summer (P<0.0001) and fall (P<0.05) (Fig. 1C). Above the age of 30 years more than 40% of the population was vitamin D-deficient, with men consistently more (P<0.05) affected than women (Fig. 1D) and in all seasons except summer (Fig. 1E). The time frame of peak SARS-CoV-2 infection in our population thus coincided with the time frame of lowest 25(OH)D levels and highest rates of vitamin D deficiency, with men (48.6%) markedly more affected than females (42.5%, P<0.05).

**Figure 1:**
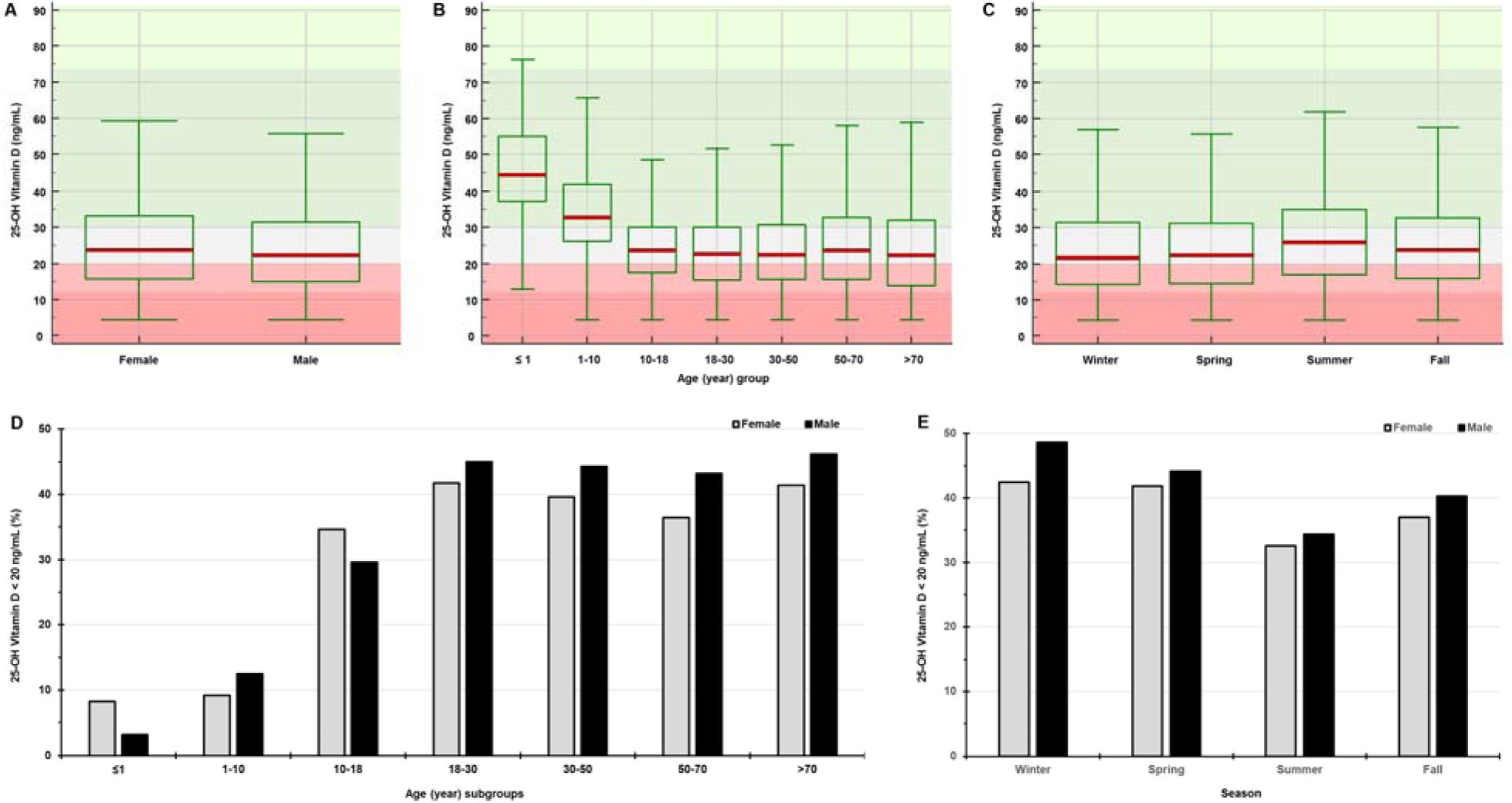
Prevalence and distribution of vitamin D deficiency in the general population as derived from 25(OH)D measurements in 16274 unselected, consecutive and unique diseased control samples Box-and-Whisker plots show median (IQR) 25(OH)D levels (ng/mL) grouped by **(A)** sex, **(B)** the indicated age groups and (C) Northern hemisphere meteorological seasons. Background color in box plots indicates normal vitamin D status (green, 25(OH)D > 30 ng/mL), vitamin D deficiency (pale red, 25(OH)D < 20 ng/mL), severe vitamin D deficiency (darker red, 25(OH)D < 12 ng/mL) and a gray zone (20 ng/mL ≤ 25(OH)D ≤ 30 ng/mL). Lower panels indicate the percentage vitamin D deficiency (25(OH)D < 20 ng/mL) in females (gray bars) and males (black bars) **(D)** in the indicated age groups (years) and **(E)** across seasons. Corresponding numbers per group, median (IQR) 25(OH)D, deficiency rates and relevant statistical differences are detailed in Table 1.

### Demographics and vitamin D status of COVID-19 patients

186 patients with PCR-confirmed SARS-CoV-2 infection were admitted to the hospital from March 1, 2020 to April 7,2020 for COVID-19 pneumonia: 109 males (median age 68 years, IQR 53-79 years) and 77 females (median age 71 years, IQR 65-74 years). 25(OH)D in COVID-19 patients was compared a control group of 2717 patients with similar age distribution, sampled in March and April, 2019. COVID-19 patients had an even lower median 25(OH)D on admission (18.6 ng/mL, IQR 12.6-25.3) than controls (21.5 ng/mL, IQR 13.9-20.8, P=0.0016) and a markedly higher percentage of vitamin D deficiency (defined as 25(OH)D < 20ng/mL): 58.6% versus 45.2% (P=0.0005) (Table 2). Considering the male preponderance in COVID-19 patients (59%) but underrepresentation in controls (37%), we then stratified 25(OH)D for sex (Fig.2A-C and Table 2). Remarkably, we observed a sexual dimorphism. Female COVID-19 patients were not more vitamin D deficient than female controls (46.8% versus 42.8%, P=0.5646, Fig.2B). Control males showed higher vitamin D deficiency rates than control females (49.2% versus 42.8%, P<0.0001 Fig.2A). In male COVID-19 patients, vitamin D deficiency was even more profound (Fig.2C), with lower median 25(OH)D (17.6 ng/mL, IQR 12.7-24.0 versus 20.3 ng/mL, IQR 13.7-28.3, P=0.0234) and a markedly higher deficiency rate (67.0% versus 49.2%, P=0.0006) than male controls.

**Figure 2:**
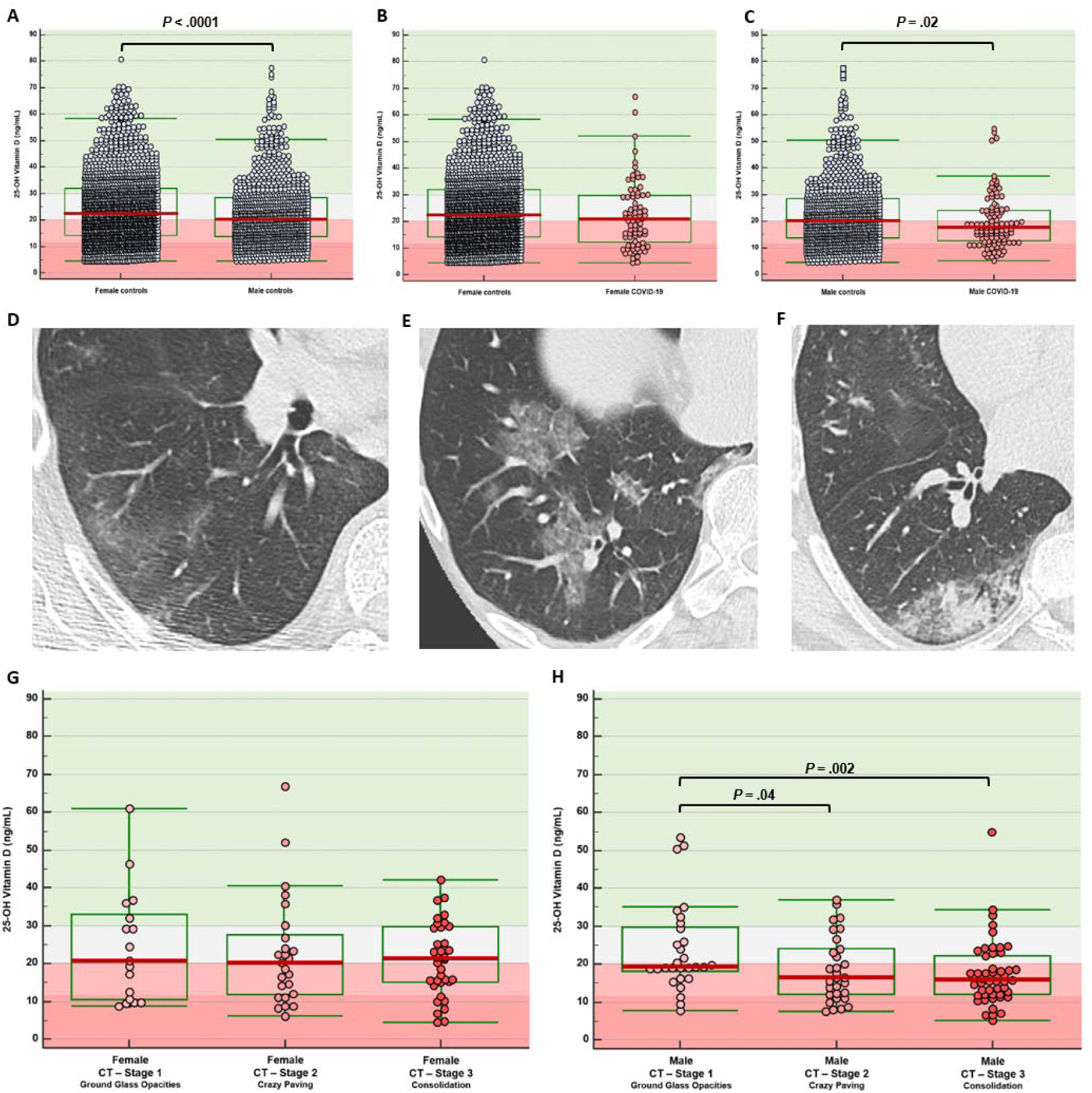
25(OH)D levels in male and female COVID-19 patients and age/season-matched controls and stratification by COVID-19 disease stage. Panels A-C: Box-and-Whisker plots showing median (red line) serum 25(OH) levels and interquartile ranges (green box) in **(A)** season- and age-matched female (n=1718) versus male (n=999) diseased controls; **(B)** female COVID-19 patients on admission (n=77) versus female controls; **(C)** male COVID-19 patients on admission (n=109) versus male controls. Panels D-F: representative images of radiological stages of COVID-19 lung disease with predominantly **(D)** ground-glass opacities in early stage; **(E)** crazy paving patterns in progressive stage 2; and **(F)** consolidation in peak stage 3. Panel G-H: box-and-whisker plots of 25(OH)D in **(G)** female COVID-19 patients and **(H)** male COVID-19 patients grouped according to radiological stage. Background color in box plots indicates normal vitamin D status (green, 25(OH)D > 30 ng/mL), vitamin D deficiency (pale red, 25(OH)D < 20 ng/mL), severe vitamin D deficiency (darker red, 25(OH)D <12 ng/mL) and a gray zone (20 ng/mL ≤ 25(OH)D ≤ 30 ng/mL). P values indicate statistical differences between groups calculated by Mann-Whitney test. Exact P values listed in Supplementary information.

### Correlation between vitamin D status and disease stage

Patients were screened by CT to determine the temporal phase of COVID-19 lung disease and classified based on the predominant radiological lesion (Fig.2D-F) as early stage 1 (ground-glass opacities), progressive stage 2 (crazy paving pattern) or peak stage 3 (consolidation). These stages are considered as proxy for the immunological phase of COVID-19 with an early phase of active viral replication in lower airways (stage 1), progressive recruitment of pro-inflammatory cells to the lung interstitial space (stage 2), ending in diffuse alveolar damage and fibrosis (stage 3). 24.7%, 29.6% and 45.7% of patients presented in stage 1, 2 and 3, respectively with similar distribution in males and females (Table 2). Analysis of 25(OH)D across radiological disease stages (Fig.2G-H, Table 2) strengthened the correlation: male COVID-19 patients showed progressively lower median 25(OH)D with advancing stage, resulting in vitamin D deficiency rates increasing from 55.2% in stage 1, 66.7% in stage 2 to 74.0% in stage 3 (P=0.0010). No such stage-dependent 25(OH)D variations were seen in female COVID-19 patients.

### Analysis of possible confounders by vitamin D-impacted comorbidities

The higher rates of vitamin D deficiency in COVID-19 patients might reflect a causal relation or be no more than a marker of poor general health or nutritional status predisposing to severe COVID-19. To identify possible confounders, we compared the prevalence of known vitamin D-impacted comorbidities such as chronic obstructive lung disease ^15,16^, coronary artery disease and diabetes, in male versus female and in vitamin D-deficient versus -replete COVID-19 patients. Prevalence of chronic lung disease, coronary artery disease and diabetes in all COVID-19 patients were 15.1%, 59.1% and 14.0%, respectively, with no differences across COVID-19 disease stages (Table 2). The high prevalence of coronary artery disease in COVID-19 patients was strongly correlated to patients’ age (r_s_ = 0.655, 95% CI 0.565-0.730, P<0.0001). Male COVID-19 patients showed similar prevalence of diabetes and coronary artery disease and a tendency towards more chronic lung diseases, that attained significance only in male COVID-19 stage 1 patients. The latter difference was, however, not related to differences in vitamin D status: male and female COVID-19 patients with normal (25(OH)D ≥ 20 ng/mL) or deficient vitamin D status showed comparable prevalence of chronic lung disease, coronary artery disease and diabetes (Table 3), indicating that the correlation between vitamin D deficiency and the risk for severe COVID-19 was not confounded by any of these comorbidities.

**Table 3:** Prevalence of comorbidities within COVID-19 patients stratified for sex and vitamin D deficiency status. Prevalence of the indicated vitamin D-impacted comorbidities are listed for all COVID-19 patients and grouped per sex (absolute number and percentage per subgroup) and compared between patients with or without vitamin D deficiency (25(OH)D < 20 ng/mL). P values calculated by chi-squared testing indicate statistical differences between prevalence of the indicated comorbidity between all, female or male vitamin D deficient versus sufficient COVID-19 patients. † Indicates no statistical differences between males and females for the indicated comorbidity.

| Patient group | Comorbidity | 25-OH-vitamin D<br>≥ 20 ng/mL | 25-OH-vitamin D<br>< 20 ng/mL | P |
| --- | --- | --- | --- | --- |
| All patients (n = 186) | n | 77 | 109 |  |
|  | Chronic lung disease, n (%) | 13 (16.9) | 15 (13.8) | .7085 |
|  | Coronary artery disease, n (%) | 48 (62.3) | 62 (56.9) | .5575 |
|  | Diabetes, n (%) | 11 (14.3) | 15 (13.8) | .9063 |
| Female patients (n = 77) | n | 41 | 36 |  |
|  | Chronic lung disease, n (%) | 4 (9.8) | 3 (8.3) | .8660 |
|  | Coronary artery disease, n (%) | 25 (61.0) | 18 (50.0) | .4594 |
|  | Diabetes, n (%) | 7 (17.1) | 4 (11.1) | .6714 |
| Male patients (n = 109) | n | 36 | 73 |  |
|  | Chronic lung disease, n (%) | 9 (25.0)<sup>†</sup> | 12 (16.4)<sup>†</sup> | .4163 |
|  | Coronary artery disease, n (%) | 23 (63.9)<sup>†</sup> | 44 (60.3)<sup>†</sup> | .8776 |
|  | Diabetes, n (%) | 4 (11.1)<sup>†</sup> | 11 (15.1)<sup>†</sup> | .7838 |

P values less than .05 were considered statistically significant.
† Additional male to female comparisons for prevalence of comorbidities showed no statistical differences (P > .05).

## Discussion

This study is one of several rapidly emerging reports that correlate vitamin D deficiency to the risk for severe presentations of COVID-19 lung disease. It is compatible with a recent study that correlated severe vitamin D deficiency in the Philippines’s population with COVID-19 disease burden and poor outcome ^9^. Our study is the first to show that more profound vitamin D deficiency predisposes to more advanced radiological stages as proxy for the severity of the inflammatory process of COVID-19 pneumonia. It also describes a remarkable sexual dimorphism, not directly explained by sex differences in overall vitamin D status in our population. This study also highlights vitamin D deficiency as a neglected health issue, even in a wealthy population with access to good nutrition and healthcare: with exception of the very young, more than 40% of our control population have 25(OH)D levels below 20 ng/mL, a widely used threshold for adequate bone health. It can be assumed that these numbers are representative for the total Belgian population that showed much higher all-cause excess mortality during the SARS-CoV-2 peak infection phase than neighboring countries according to the European Mortality Monitoring project ^17^ despite largely sufficient hospital and ICU capacity. Our data thus represent a first field validation of a recent global epidemiological analysis that clearly correlated the overall incidence and mortality of COVID-19 to geographical latitude, nations’ nutritional status and the associated known impact on vitamin D deficiency as proposed causal factor^13^.

Vitamin D deficiency might be cause or consequence of severe COVID-19 lung disease. It might also be simply a surrogate marker for an underlying confounding comorbidity or general indicator of poor nutrition and ill health. Chronic obstructive pulmonary disease and asthma have been shown to alter 25(OH) metabolism or sequestration ^15,16^ with lower increments in circulating 25(OH)D status after controlled repletion. Vitamin D deficiency has also been correlated ^18^ to known COVID-19 risk factors such as hypertension and diabetes. Our analysis of comorbidities strongly argue against such confounding effect: vitamin D deficiency was strongly correlated to the risk of advanced COVID-19, specifically in males, but vitamin D deficient COVID-19 patients did not show higher prevalence of known vitamin D-impacted diseases such as diabetes, coronary artery disease and chronic lung disease. Another possible confounder is increased catabolism of circulating 25(OH)D in COVID-19 patients, attributed to extrarenal CYP27B1 activity and VDR expressed by the expanded repertoire of immune cells. In a USA population survey, subjects with CRP>5 mg/L showed 1.6 ng/L lower median 25(OH)D ^20^. However, the stable 25(OH)D levels in female COVID-19 patients across all stages of the disease argue against such effect. Combined, our data thus support a causal role of vitamin D deficiency, in line with a recent modeling study using causal interference analysis of global COVID-19 incidence and mortality data ^13^.

A substantial body of evidence explains a causal pathophysiological role of vitamin D in the severity of respiratory viral infections. The immunological response to SARS-CoV-2 shows many similarities to the response to SARS-CoV ^21^: in individuals with delayed type I/III interferon response and delayed viral clearance, progressive recruitment of neutrophils and pro-inflammatory Th1/M1-polarized immune cells contribute to endothelial and alveolar cell death and in some patients trigger a cytokine storm ^1^ that enhances diffuse alveolar damage. Severe COVID-19 can thus be conceptualized as an unbalance between pro-inflammatory type I immune response required for viral clearance and tolerogenic type II response required for repair ^1,2,21^. Vitamin D modulates the immunological response to respiratory viruses at various phases: in the early phase of infection it limits viral entry and replication by boosting cathelicidins/defensins expression in respiratory epithelia ^5^. Later on it exerts a tolerogenic effect by directly mediating IL-4/IL-13-dependent polarization towards M2-macrophages and Th2 CD4 T cells ^6^. The mammalian immune system shows conserved estrogen/androgen-dependent sexual dimorphism ^22^. SARS-CoV-infected female mice show lower viral replication, lower recruitment of inflammatory monocytes and neutrophils to the lungs and less alveolar and endothelial cell death ^23^. If vitamin D deficiency favors a pro-inflammatory balance, the higher rates of vitamin D deficiency in male humans might thus act in concert with estrogen/androgen-dependent immune differences and contribute to higher incidence and severity in male COVID-19 patients.

Several randomized controlled interventional trials with vitamin D supplementation for bone health or to reduce mortality in cardiovascular disease and cancer gave mixed or negative results, entailing a sense of indifference from the medical community towards the value of vitamin D monitoring and supplementation. An overlooked issue, however, was that many of those interventional trials showed inappropriate design by targeting populations with no prior vitamin D deficiency, and were not guided by actual 25(OH)D measurements ^24^. A prior meta-analysis indicated that vitamin D supplementation reduces the incidence and severity of acute respiratory infections but only in those patients with a deficient vitamin D level at the start as measured by 25(OH)D ^25^.

In conclusion, our study shows a strong correlation between vitamin D deficiency and severe COVID-19 lung disease that is not explained by confounding comorbidities. In light of the established immunological functions of vitamin D, it is compatible with a causal role of vitamin D deficiency that can explain variations in disease burden of COVID-19 across geographies, skin pigment types (African/Latino), body mass (obese), socioeconomic status (poor) and lifestyle (institutionalized people). Given the global prevalence of vitamin D deficiency, our data thus argue for vitamin D supplementation as inexpensive, safe and readily available mitigation of the syndemic convergence of two pandemics.

## Contributor and guarantor information

D.D.S., K.D.S and G.M. were responsible for planning and design of the study. Statistical analysis was done by D.D.S. P.H. was responsible for laboratory analyses. S.G. and K.D.S. were responsible for CT and structured reporting of radiological data. All authors contributed to the manuscript preparation, read and approved it. G.M. was responsible for preparing the manuscript and is guarantor of the study. The guarantor affirms that the manuscript is an honest, accurate, and transparent account of the study being reported; that no important aspects of the study have been omitted; and that any discrepancies from the study as planned (and, if relevant, registered) have been explained. The authors declare that they will share anonymized source data of this study upon written request to the guarantor.

## Data Availability

All data required for independent reproduction of our study are included in the manuscript and online supplementary files. The raw data are freely available on email request.

